# Prediction of a major adverse coronary event in Women through CORSWO

**DOI:** 10.1101/2023.03.22.23287603

**Authors:** Guillermo Romero-Farina, Santiago Aguadé-Bruix, Ignacio Ferreira-González

## Abstract

**BACKGROUND:** In women, risk stratification for a major adverse coronary event (MACE) is complex, and moreover women have often been underrepresented in cardiovascular studies. This study aims to establish a **CO**ronary **R**isk **S**core in **WO**men (CORSWO) to predict MACE.

**METHODS:** From a cohort of 25,943 consecutive patients referred for clinical gSPECT-MPI (gated single-photon emission computed tomography myocardial perfusion imaging), 2,226 women (aged 66.7±11.6 years) were included. During the follow-up (mean 4±2.7 years) post gSPECT-MPI, MACE (unstable-angina requiring hospitalization, non-fetal myocardial infarction, coronary revascularization, cardiac death) was assessed. The patients were divided into training (n=1460) and validation (n=766) groups. To obtain the predictor model, LASSO-regression analysis with 10-fold cross-validation was used.

**RESULTS:** In training group, 148 women had MACE (0.026/patient/year). The best model (ROC area:0.8, Brier score:0.0777) to stratify women included: age >69 years (OR:1.6); diabetes mellitus (OR:2); angina-history (OR:1.6); nitrate (OR:1.5); left bundle branch block (OR:1.2); pharmacological test (OR:1.6); ST-segment-depression (≥1mm) (OR:2); stress angina (OR:1.3); myocardial ischemia >5% (OR:2.6); perfusion defect at rest >9% (OR:2.4); perfusion defect at stress >6% (OR:1.7); end-systolic volume index >15 ml (OR:1.6); and left ventricular ejection fraction <50% (OR:1.2). This model was validated (validation group) with a strong prediction (ROC area:0.8, Brier score:0.0747). The CORSWO obtained from these variables allows the stratification of women into five risk levels: very low (score:0,HR:1), low (score:1-2,HR:1.5), moderate (score:3-6,HR:2.7), high (score:7-10,HR:6.9) and very high (score:≥11,HR:21.7).

**CONCLUSIONS:** In a clinical practice setting we can obtain an excellent coronary risk stratification in women, however at the expense of multiple variables.

**CLINICAL PERSPECTIVE:** *What Is New?:* The coronary risk stratification of women depends on of the multiple clinical, exercise and imaging variables. This new risk score allows the risk to be calculated for individual women in a simple way with a mean of a 4-year follow-up.

*What Are the Clinical Implications?:* CORSWO is an effective tool to stratify the risk for major adverse coronary event in 5 risk levels, very low, low, moderate, high and very high risk with a good accuracy.

## INTRODUCTION

Coronary artery disease (CAD) continues to be the leading cause of death among women in the United States^1^, Latin America^2^, Europe^3^, Asia^4^ and in the Pacific countries^5, 6^. Despite all the studies carried out in relation to coronary risk stratification in women with an exercise treadmill test^7^, isotopic techniques^8–13^, cardiac magnetic resonance^14^, coronary computed tomography angiography^15, 16^, and stress echocardiography^17^, the assessment of CAD continues to be a challenge.

The presentation of CAD in women is 7-10 years later when compared to men and with atypical symptoms^18, 19^. Moreover, women are more likely to present acute myocardial infarction without chest pain and have a higher mortality rate than men, especially among the younger age groups^19–21^. On the other hand, myocardial infarction with epicardial coronary stenosis is the most important manifestation of CAD in men. However, women can present myocardial ischemia with no obstructive CAD caused by microvascular dysfunction, vasospastic disorders, or any other mechanisms such as spontaneous coronary artery dissection^22^.

Another reality is that women have often been underrepresented in cardiovascular studies, and few studies in cardiovascular medicine have addressed the issue of coronary risk focused on clinical and imaging outcomes in women alone^8–17^. Furthermore, among women, there exists an increase in the impact of traditional atherosclerotic cardiovascular disease risk factors including diabetes mellitus, hypertension, dyslipidemia, smoking, obesity, and physical inactivity^1^. Recently, an AHA presidential advisory was published, which explains the critical need for research toward achieving significant progress in the health and well-being of all women^22^. For all these reasons, it is important to assess the coronary risk associated with imaging techniques. Thus, from a clinical point of view the risk stratification by means of clinical variables and non-invasive techniques is important and necessary. Therefore, this study aims to establish a **CO**ronary **R**isk **S**core in **WO**men (CORSWO) to predict MACE in a large population-based series of women **(Central illustration)**.

## METHODS

### Study population

This study complies with the Declaration of Helsinki and was reviewed and approved by the Ethics Committee of Vall d’Hebron Hospital (registered as PR(AG)168.2012). A written consent was obtained from all patients prior to the gSPECT-MPI.

This observational cohort study is conducted in a University Reference Hospital. Between 2000 and 2018, of 25,943 consecutive patients who underwent gated SPECT myocardial perfusion imaging (gSPECT-MPI), 2,226 women were included **(Figure 1)**. This study is a retrospective analysis of data extracted from a prospectively collected database. Every woman included is between 40 and 93 years of age. All the women (n= 2,226) had been referred to our Nuclear Cardiology Unit for risk stratification or evaluation of their disease.

**Figure 1.**
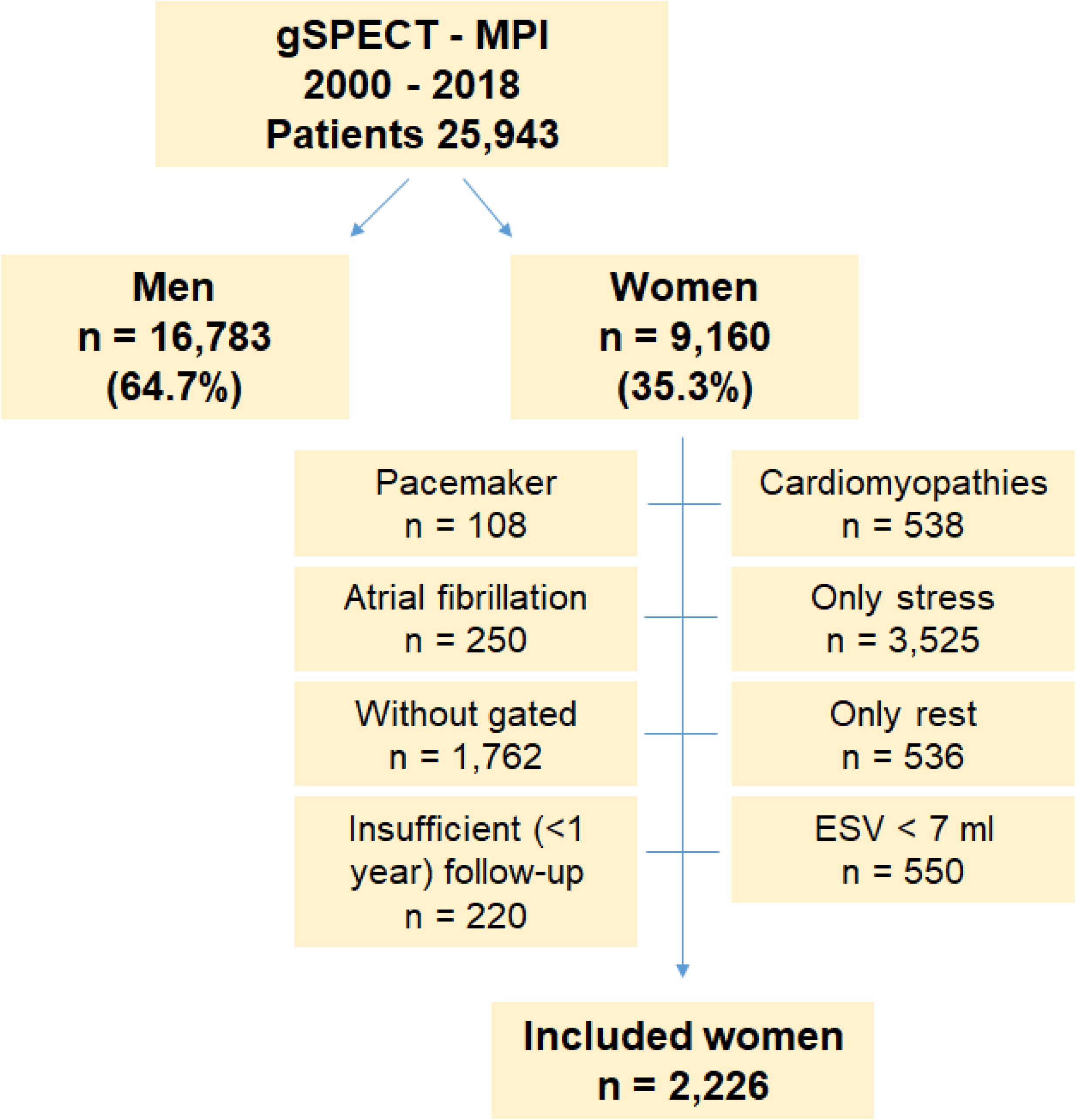
Flow diagram showing women included in the study. ESV, end-systolic volume.

### Gated SPECT myocardial perfusion imaging

All 2226 patients underwent stress-rest gSPECT-MPI: 1410 patients had a subjective maximal exercise myocardial perfusion imaging; 407 patients had submaximal exercise plus pharmacological test, due to their inability to achieve 5 METs or 80 % of predicted peak heart rate for their age; and 409 patients had only pharmacological myocardial perfusion imaging because they were unable to exercise due to very low functional capacity.

For stress-rest gSPECT-MPI studies, a short, 1-day stress-rest protocol with 99mTc-tetrofosmin was adopted. A first dose of 296 MBq was administered 30-60 seconds before the end of the stress test, and a second 814 MBq dose was administered at rest, the two separated by an interval of over 45 minutes. In patients with a body mass index ≥30 (Obesity, n=183 [27.2%]), a 2-day protocol was adopted, administering the same dose (814 MBq) for stress and rest. For the one-day protocol, the image acquisition time was about 18 minutes for the gated stress images and about 14 minutes for the gated rest images, to compensate for the low-dose stress and high-dose rest differences. In the 2-day protocol, the image acquisition time was 14 minutes for both stress- and rest-gated images. CT attenuation correction was performed when the nuclear cardiologist considered it necessary.

A scintigraphic perfusion score ranging from 0 to 4 was assigned to each of the 17 segments in both stress (SSS, summed stress score) and rest (SRS, summed rest score) (0= normal perfusion, 1=mild hypoperfusion, 2=moderate hypoperfusion, 3=severe hypoperfusion, and 4=no uptake). The summed difference score (SDS) was calculated from the sum of the segmental difference scores between stress and rest scans. SSS, SRS, and SDS were converted to percent total myocardium (SSS%, SRS%, SDS%) by means of the following formula: summed scores / 68 (maximum potential score = 4 × 17) × 100. The calculation of the left ventricular ejection fraction (LVEF), end-diastolic volume (EDV) index, and end-systolic volume (ESV) index were performed automatically in rest gSPECT-MPI by means of the automatic delineation of the endocardial and epicardial borders using the quantitative QGS-software package (Cedars-Sinai Medical Center, Los Angeles, CA).

### Clinical outcomes

The minimum follow-up time was 1 year (maximum 10 years), and the mean follow-up was 4 ± 2.7 years (median 3.5 years, 25th-75th percentile: 1.4-6). All patients were followed up in the hospital, and the follow-up information was obtained by clinical history in. There were no missing patients during the follow-up. All patients were followed up for MACE (UA, unstable angina; non-fatal MI, non-fatal myocardial infarction; CR, coronary revascularization; and CD, cardiac death). MI was defined according to the new 2003 and 2005 definitions that included the use of troponins^23, 24^, and UA according to the guideline of 2010^25^. The primary endpoint was the first occurrence of a composite of UA, non-fatal MI, CR and CD (MACE).

### Statistical analysis

All continuous data were expressed as mean (SD, standard deviation) and all non-continuous variables were expressed as percentages. Continuous variables were compared using the Student *t* test for unpaired samples. Differences between proportions were compared using the χ2 test. Fisher’s exact test was used when <5 patients were expected in any subgroup.

Variables for multivariate analysis were selected when they presented a *p* ≤ 0.05 in the univariate analysis or when they were considered of clinical relevance.

The predictive model to MACE was constructed using LASSO (Least Absolute Shrinkage and Selection Operator) regression analysis with 10-fold cross-validation. The population was divided in a training group (65.6%, n=1460) and a validation group (34%, n=766).

According to the model obtained, the probability of MACE was calculated for every patient in the training group. The probability of MACE in the validation group was calculated by means of the coefficients obtained in the training group.

Moreover, a BRIER score was calculated in the training and validation groups. The Brier score measures the total difference between the event and the forecast probability of that event as an average squared difference. As a benchmark, a perfect forecaster would have a Brier score of 0, a perfect misforecaster would have a Brier score of 1^26^.

Then, in training group, a z-score was calculated for every prognostic variable; and also a five risk level of z-score was obtained according to MACE/patient/year: very low risk (VLR, < 0.7 MACE/year), low risk (LR, 0.7 to 1 MACE/year), moderate risk (MR, 1 to 3 MACE/year), high risk ( HR, 3 to 5 MACE/year), and very high risk (VHR, > 5 MACE/year).

This z-score was applied in the validation group, and the MACE/patient/year ratio of the 5-risk levels were compared between the testing and validation groups. No missing data existed for the variables included in the prognostic score.

To compare the survival curves of the different risk levels, Cox regression analysis was used. Hazards ratios (HR), 95% confidence intervals (CIs), and statistical significance for each group in the model was determined. We tested the proportional hazards assumption in all Cox regression. We considered that there is no evidence that our specification violates the proportional hazards assumption when *P* value >0.05, and when the plotted curves were parallel.

All statistical tests were two-sided. A value of *P* value <0.05 was considered to be indicative of statistical significance. Data was analysed by STATA 17 (StataCorp, College Station, TX, USA).

## RESULTS

During the follow-up (4 ± 2.7 years), the overall mortality and MACE in all cohort (n=2226) was 6.7% (n=218; 0.024/patient/year) and 6.6% (n=220; 0.025/patient/year) respectively. UA: 2.6% (n=57; 0.0067/patient/year); non-fatal MI: 1.7% (n=38; 0.0044/patient/year); CR: 2.9% (n=64; 0.0075/patient/year), and CD: 4.3% (n=95; 0.0111/patient/year). Of the 64 patients who underwent CR, in 33 (50.8%) patients the CR was early (≤ 3 months post gSPECT-MPI) and in 32 (49.2%) patients the CR was late.

### Training group

Baseline characteristics, in the training group according to all-cause mortality and MACE, are shown in **Table 1**. The overall mortality and MACE in training group (n=1460) were 10.3% (n=150; 0.025/patient/year) and 10.1% (n=148; 0.026/patient/year) respectively. For the multivariate LASSO regression analysis 19 variables were included (age > 69 years, diabetes, hypertension, hypercholesterolemia, current smoking, left bundle branch block (LBBB), previous myocardial infarction, previous coronary revascularization, angina, nitrate, stress angina, pharmacological test, ↓ ST mm ≥ 1, SRS > 9.6, SSS > 6.6, SDS > 5.2, ESV index > 15 ml, EDV index > 38 ml, and LVEF < 50%). The obtained predictive model to MACE is shown in **Table 2**. **Figures S1 and S2** show the internal validation of the predictor model. **Figure 2** shows the analysis of Brier score. Then, the z-score (from 0 to 28) was calculated for every predictive variable **(Table 2)**. According to MACE/patient/year different levels of z-score risk were calculated **(Table 3)**. **Figure 3** shows the relationship between the predicted (CORSWO: VLR, LR, MR, HR and VHR) and the observed (MACE). A comparison by means of Cox regression analysis between patients with different risk levels and MACE was made **(Figure 4A)** (Harrell’s c-index: 0.8).

**Figure 2.**
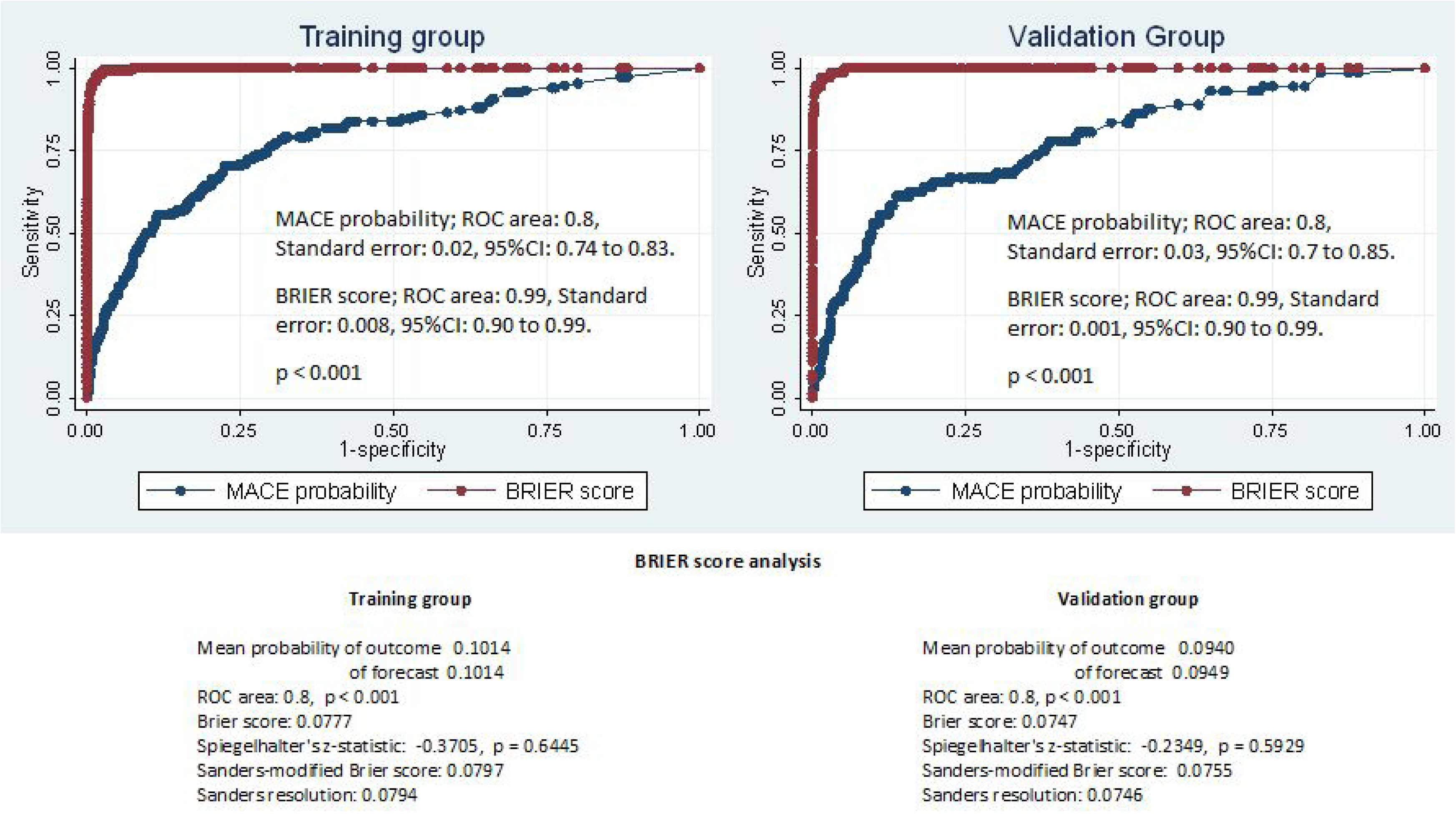
ROC curve analysis to evaluate the ability of the final model to predict MACE; shows an excellent predictive value.

**Figure 3.**
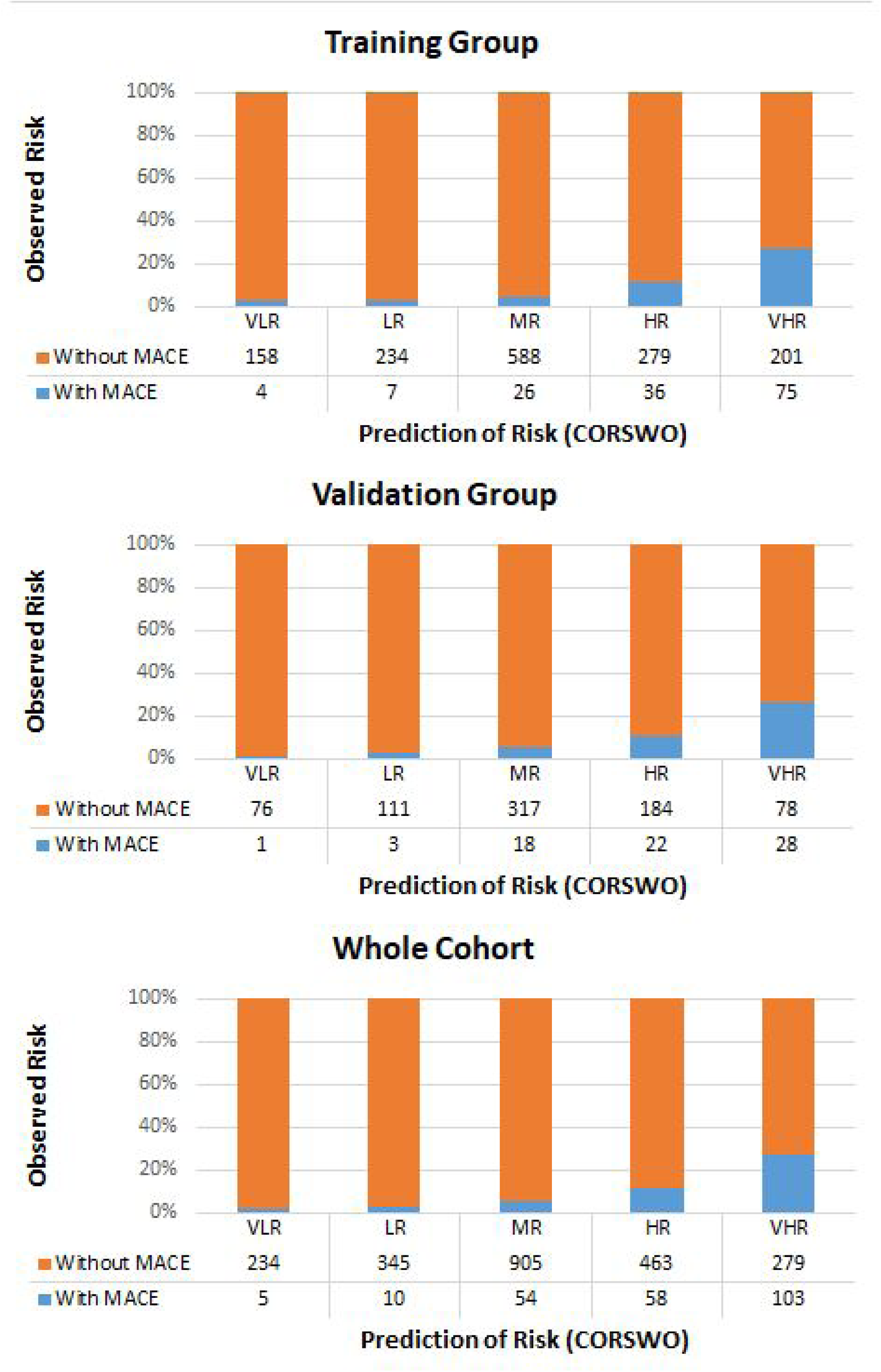
Coronary Risk Score in Women (CORSWO) to predict MACE. When increasing the levels of CORSWO (Prediction of risk) calculated by using a z-score, the prevalence of MACE (observed risk) increases too. HR, high risk; LR, low risk; MACE, major adverse coronary event; MR, moderate risk; VHR, very high risk; VLR, very low risk.

**Figure 4.**
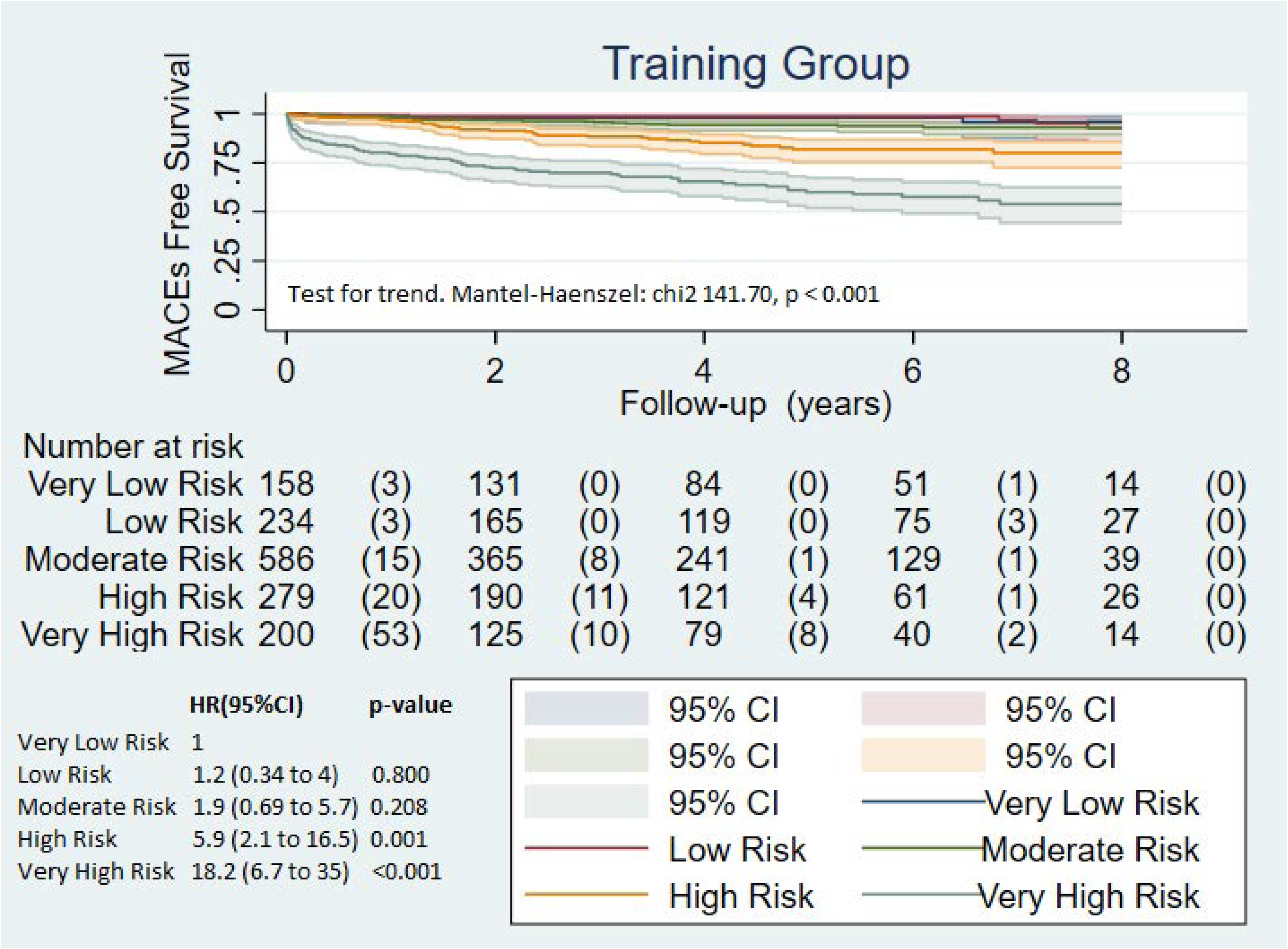
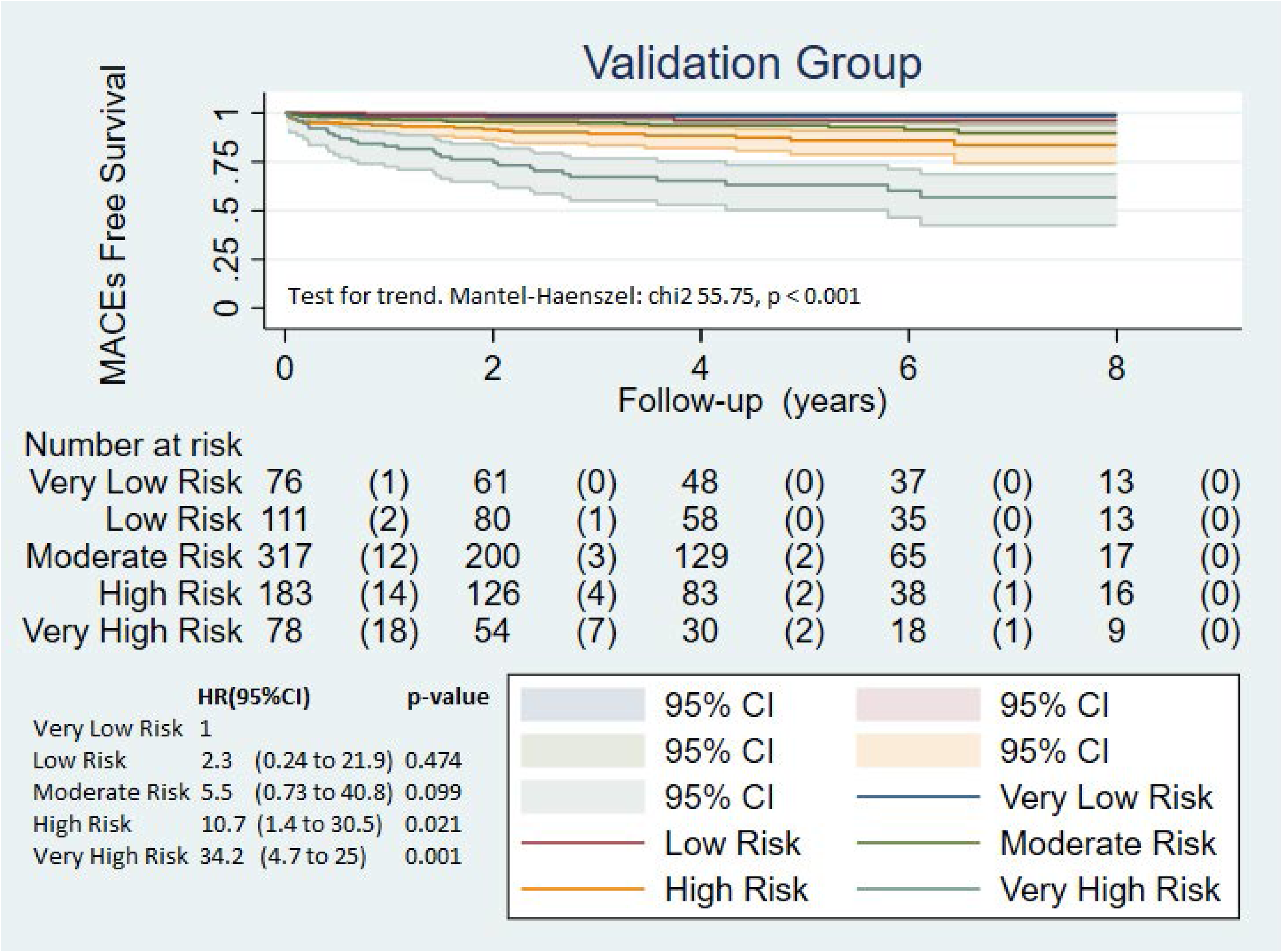
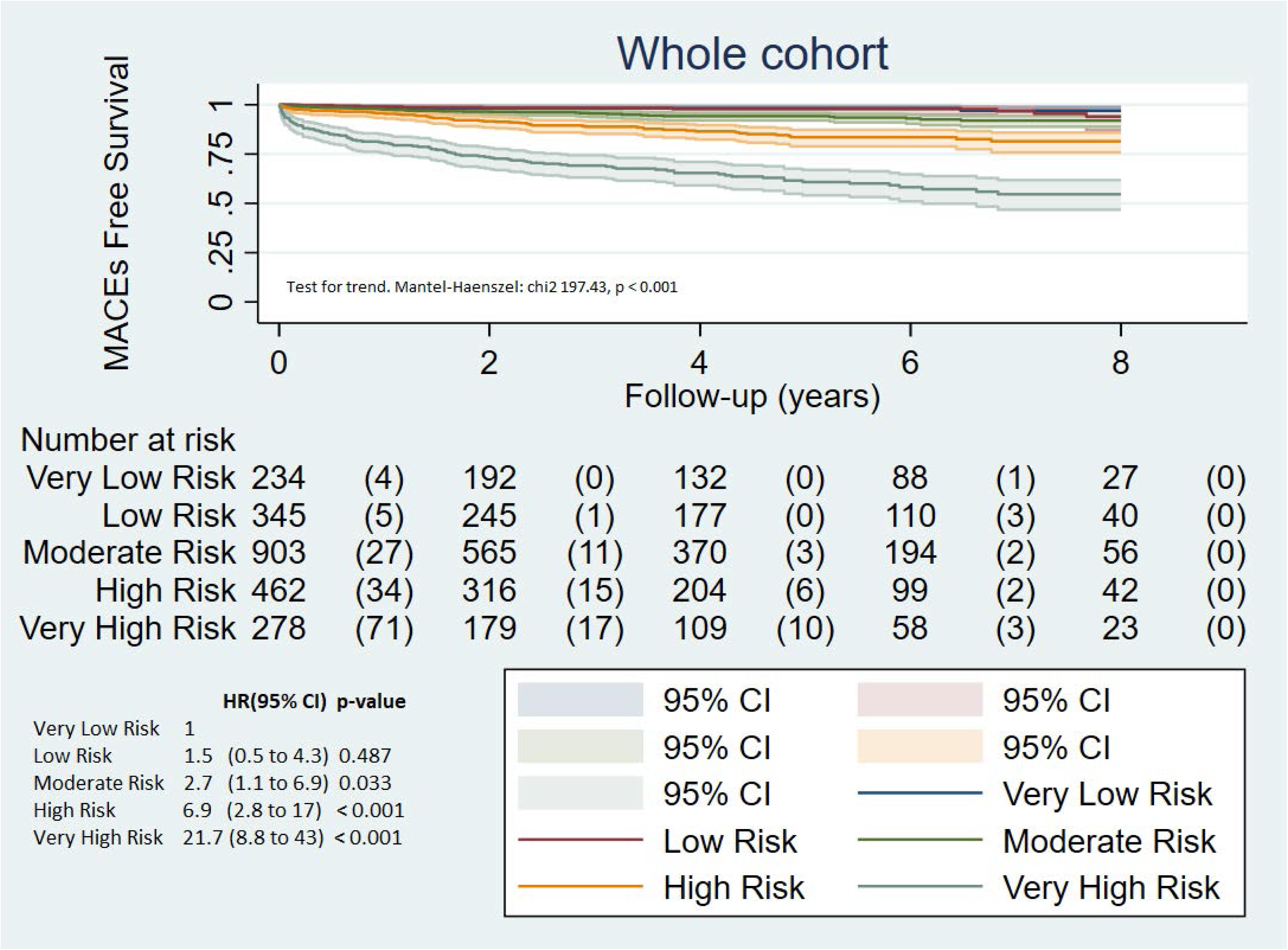
Kaplan-Meier curve analysis. During follow-up post gSPECT-MPI, for a one-unit increase in risk revel the rate ratio to MACE increases 2.3 (95%CI: 2.01 to 2.65) in training group, 2.2 (95%CI: 1.8 to 2.7) in validation group, and 2.38 (95%CI: 2.02 to 2.54) in whole cohort. CI, confidence interval; HR, hazard ratio.

**Table 1.** Baseline **c**linical, stress test, and gated SPECT characteristics of the training group according to all-cause mortality and MACE.

|  | **Overall**<br>**(n=1460)** | **Alive**<br>**(n=1310)** | **All-cause**<br>**mortality**<br>**(n=150)** | ***P*-value** | **Without**<br>**MACE**<br>**(n= 1312)** | **With**<br>**MACE**<br>**(n=148)** | ***P*-value** |
| --- | --- | --- | --- | --- | --- | --- | --- |
| **Demographics** |  |  |  |  |  |  |  |
| Age (years), mean (SD) | 66.7 (11.9) | 66.1 (11.8) | 72.7 (11.1) | < 0.001 | 66.3 (11.9) | 70.8 (10.9) | <0.001 |
| Age > 69 years, *n* (%) | 733 (50.21) | 621 (47.4) | 112 (74.7) | <0.001 | 639 (48.7) | 94 (63.5) | 0.001 |
| Body surface (m<sup>2</sup>), mean (SD) | 1.8 (0.17) | 1.8 (0.18) | 1.8 (0.17) | 0.986 | 1.8 (0.2) | 1.7 (1.6) | 0.994 |
| Body mass index (kg/m<sup>2</sup>), mean (SD) | 27.8 (4.6) | 27.8 (4.7) | 27. (1 3.9) | 0.105 | 27.8 (4.4) | 27.8 (5.9) | 0.960 |
| Obesity (BMI ≥ 30), *n* (%) | 340 (23.3) | 309 (23.4) | 31 (20.7) | 0.423 | 305 (23.3) | 35 (23.7) | 0.913 |
| **Medical history** |  |  |  |  |  |  |  |
| Hypertension, *n* (%) | 964 (66.03) | 855 (65.3) | 109 (72.7) | 0.070 | 855 (65.17) | 109 (73.7) | 0.039 |
| Hypercholesterolemia, *n* (%) | 748 (51.2) | 680 (51.9) | 68 (45.3) | 0.127 | 663 (50.5) | 85 (57.4) | 0.111 |
| Smoking, *n* (%) | 188 (12.9) | 177 (13.5) | 11 (7.3) | 0.032 | 171 (13) | 17 (11.5) | 0.594 |
| Diabetes mellitus, *n* (%) | 329 (22.5) | 257 (19.6) | 72 (48) | < 0.001 | 265 (20.2) | 64 (43.2) | < 0.001 |
| CRF ≥ 1 (%) | 1224 (83.8) | 1094 (83.5) | 130 (86.7) | 0.320 | 1090 (83.1) | 134 (90.5) | 0.019 |
| Previous MI, *n* (%) | 303 (20.8) | 261 (19.9) | 42 (28) | 0.021 | 251 (19.1) | 52 (35.1) | < 0.001 |
| Previous CR, *n* (%) | 184 (12.6) | 164 (12.5) | 20 (13.3) | 0.776 | 158 (12.04) | 26 (17.6) | 0.055 |
| **Electrocardiogram** |  |  |  |  |  |  |  |
| Sinus rhythm, *n* (%) | 1460 (100) | 1310 (100) | 150 (100) | - | 1312 (100) | 148 (100) | - |
| Left bundle branch block, *n* (%) | 109 (7.5) | 84 (6.4) | 25 (16.7) | < 0.001 | 89 (6.8) | 20 (13.5) | 0.007 |
| **Symptoms at gated SPECT** |  |  |  |  |  |  |  |
| Angina, *n* (%) | 255 (17.5) | 218 (16.6) | 37 (24.7) | 0.014 | 211 (16.1) | 44 (29.7) | < 0.001 |
| **Medical treatment at gated SPECT** |  |  |  |  |  |  |  |
| Beta-blocker, *n* (%) | 500 (34.3) | 438 (33.4) | 62 (41.3) | 0.053 | 425 (32.4) | 75 (50.7) | < 0.001 |
| Nitrate, *n* (%) | 422 (28.9) | 373 (28.7) | 49 (32.7) | 0.283 | 355 (27) | 67 (45) | < 0.001 |
| Calcium-channel blocker, *n* (%) | 246 (16.9) | 217 (16.6) | 29 (19.3) | 0.391 | 216 (16.5) | 30 (20.3) | 0.241 |
| **Stress test** |  |  |  |  |  |  |  |
| Exercise test, *n* (%) | 923 (63.2) | 845 (64.5) | 78 (52) | 0.003 | 846 (64.5) | 77 (52) | 0.003 |
| Exercise test plus pharmacological test, *n* (%) | 275 (18.8) | 262 (20) | 13 (8.7) | < 0.001 | 249 (18.9) | 26 (17.6) | 0.677 |
| Pharmacological test, *n* (%) | 262 (17.9) | 203 (15.5) | 59 (39.3) | < 0.001 | 217 (16.5) | 45 (30.4) | < 0.001 |
| Stress angina, *n* (%) | 179 (12.3) | 169 (12.9) | 10 (6.7) | 0.025 | 155 (11.8) | 24 (16.2) | 0.122 |
| ↓ ST mm ≥ 1, *n* (%) | 213 (14.6) | 194 (14.8) | 19 (12.7) | 0.481 | 176 (13.4) | 37 (25) | < 0.001 |
| **SPECT** |  |  |  |  |  |  |  |
| SRS, mean (SD) | 1.4 (3.6) | 1.2 (3.2) | 3.5 (5.8) | < 0.001 | 1.1 (3.1) | 3.9 (61) | < 0.001 |
| SRS% > 9.6, *n* (%) | 90 (6.2) | 64 (4.9) | 26 (17.3) | < 0.001 | 115 (8.4) | 33 (36.7) | < 0.001 |
| SSS, mean (SD) | 2.4 (4.5) | 2.1 (4) | 5.4 (6.7) | < 0.001 | 1.9 (3.7) | 6.7 (7.4) | < 0.001 |
| SSS% > 6.6, *n* (%) | 150 (10.3) | 84 (7.1) | 66 (23.8) | < 0.001 | 70 (5.9) | 78 (28.2) | < 0.001 |
| SDS, mean (SD) | 1 (2.3) | 0.9 (2.1) | 2 (3.4) | < 0.001 | 0.8 (1.7) | 2.8 (4.5) | < 0.001 |
| SDS% > 5.2, *n* (%) | 150 (10.3) | 120 (9.1) | 30 (17.1%) | < 0.001 | 120 (9.1) | 30 (20.7) | < 0.001 |
| **Gated** |  |  |  |  |  |  |  |
| EDV (ml), mean (SD) | 81.3 (44.9) | 79.6 (3.8) | 95.8 (51) | < 0.001 | 79.8 (42.7) | 94.7 (59.1) | < 0.001 |
| EDV index (ml/m<sup>2</sup>), mean (SD) | 45.5 (28.9) | 44.4 (25.1) | 54.5 (30.3) | < 0.001 | 44.5 (24.7) | 53.7 (33.6) | < 0.001 |
| EDV index > 38 ml (ml/m<sup>2</sup>), *n* (%) | 705 (48.3) | 612 (46.7) | 93 (62) | < 0.001 | 616 (46.9) | 89 (60.1) | 0.002 |
| ESV (ml), mean (SD) | 37.2 (38.5) | 35.4 (37.4) | 52.4 (44.4) | < 0.001 | 35.6 (36.4) | 51.5 (52) | < 0.001 |
| ESV index (ml/m<sup>2</sup>), mean (SD) | 20.8 (21.9) | 19.8 (21.1) | 29.9 (26.3) | < 0.001 | 19.9 (20.7) | 29.3 (29.3) | < 0.001 |
| ESV index > 15 ml (ml/m<sup>2</sup>), *n* (%) | 643 (44) | 550 (41.9) | 93 (62) | < 0.001 | 547 (41.7) | 96 (64.8) | <0.001 |
| LVEF (%), mean (SD) | 60.8 (16.1) | 61.9 (15.5) | 51.3 (17.8) | < 0.001 | 61.8 (15.7) | 52.4 (16.7) | < 0.001 |
| LVEF < 50%, *n* (%) | 690 (47.3) | 592 (45.2) | 98 (65.3) | < 0.001 | 591 (45) | 99 (66.9) | < 0.001 |

CRF, coronary risk factors; EDV, end-diastolic volume; ESV, end-systolic volume; LVEF, left ventricular ejection fraction; MI, myocardial infarction; CR, coronary revascularization; SD, standard deviation; SDS, summed difference score; SDS%, ischemic myocardium percentage; SMI, silent myocardial ischemia; SRS, summed rest score; RSR%, necrotic myocardium percentage; SSS, summed stress score; SSS%, percentage of myocardium with altered stress perfusion. Cut-off values for quantitative variables were obtained by ROC curve analysis.

**Table 2.** Predicted model obtained to create CORSWO.

| **MACE predictor model in women** | **Coefficients** | **OR (95% CI)** | **Z-score** |
| --- | --- | --- | --- |
| Constant | -3.509891 |  |  |
| Age > 69 years | 0.2660845 | 1.6 (0.99 to 2.2) | 2 |
| Diabetes | 0.7044666 | 2 (1.3 to 2.9) | 3 |
| Left bundle branch block | 0.0722992 | 1.2 (0.7 to 2.2) | 1 |
| Angina history | 0.3505415 | 1.6 (0.9 to 2.7) | 2 |
| Nitrate | 0.3088062 | 1.5 (0.9 to 2.2) | 2 |
| Pharmacological stress | 0.3204405 | 1.6 (1.1 to 2.5) | 2 |
| ↓ ST mm ≥ 1 | 0.5171864 | 2 (1.2 to 3.2) | 3 |
| Stress angina | 0.1207119 | 1.3 (0.8 to 2.3) | 1 |
| Myocardial necrosis > 9.6% | 0.7880286 | 2.4 (1.2 to 4.7) | 3 |
| Myocardial ischemia > 5.2% | 0.8685052 | 2.6 (1.5 to 4.5) | 4 |
| Myocardial perfusion stress > 6.6% | 0.6085073 | 1.7 (0.9 to 3.1) | 2 |
| ESV-index > 15 ml. | 0.3412926 | 1.6 (0.9 to 2.6) | 2 |
| FEVI < 50% | 0.1078567 | 1.1 (0.7 to 2) | 1 |
| Total Z-score |  |  | 28 |

CORSWO, COronary Risk Score in WOmen; ESV, end-systolic volume; LVEF, left ventricular ejection fraction.

**Table 3.**
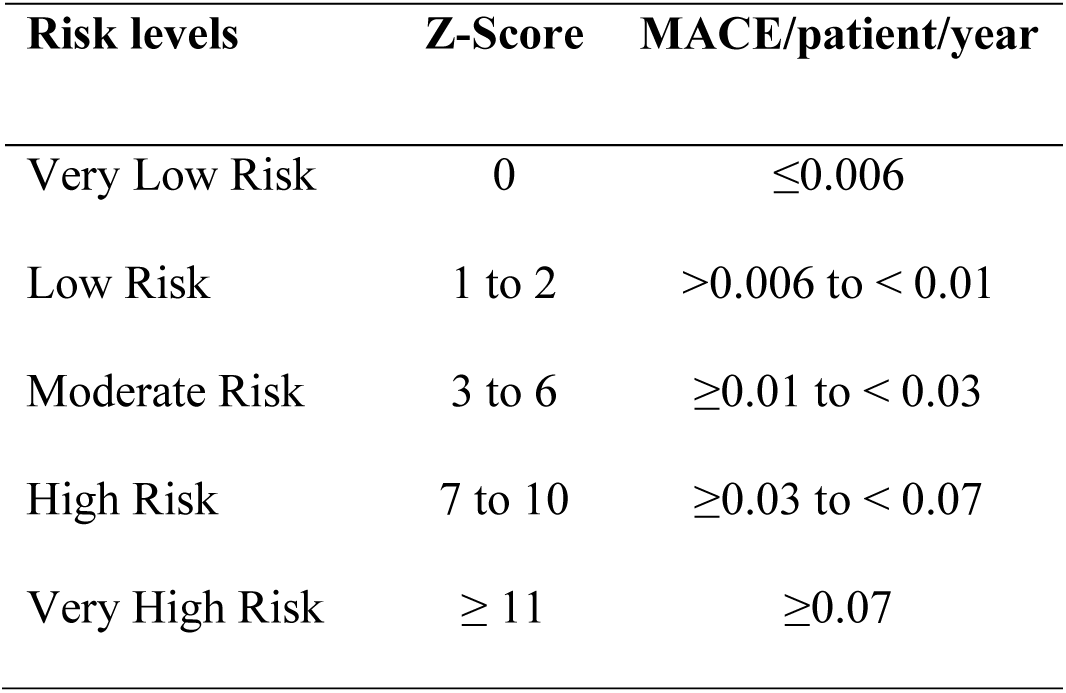
The Z-score in training group according to MACE/patient/year.

### Validation group

Baseline characteristics in training group according to all-cause mortality and MACE are shown in **Table 4**. The overall mortality and MACE in validation group (n=766) were 8.9% (n=68; 0.022/patient/year) and 9.4% (n=72; 0.024/patient/year) respectively. To validate CORSWO in women for MACE, we used the predictive variables obtained in the training group **(Table 2)**: age > 69 years, diabetes, left bundle branch block, angina history, nitrate treatment, pharmacological test, , ↓ ST mm ≥ 1, stress angina, SRS > 9.6, SDS > 5.2, SSS > 6.6, ESV index > 15 ml, LVEF < 50%. We obtained the prognosis index using the following regression equation: −3.509891 + (age > 69 years × 0.2660845) + (diabetes × 0.7044666) + (left bundle branch block × 0.0722992), + (angina history × 0.3505415) + (nitrate treatment × 0.3088062) + (pharmacological test × 0.3204405) + (↓ ST mm ≥ 1 × 0.5171864) + (stress angina × 0.1207119) + (SRS > 9.6 × 0.7880286) + (SDS > 5.2 × 0.8685052) + (SSS > 6.6 × 0.6085073) + (ESV index > 15 ml × 0.3412926) + (LVEF < 50% × 0.1078567). Then, we calculate the probability of MACEs = 1 / (1 + Exp [-(prognosis index)]). **Figure 2** shows the ROC curve, and the Brier score analysis. The z-score obtained in training group **(Table 2)** was applied in every variable to obtain different levels of risk according to the range of z-score **(Table 3)**. A comparison of MACE/patient/year between the risk levels of training and validation group was made **(Table S1)**. Furthermore, a comparison by means of Cox regression analysis between patients with different risk levels and MACEs was made in validation group **(Figure 4B)** (Harrell’s c-index: 0.8), and in all cohort (n=2226) **(Figure 4C)**. When increasing the levels of CORSWO (Prediction of risk) calculated by using a z-score, the prevalence of MACE (observed risk) increases too **(Figure 3)**.

**Table 4.** Baseline **c**linical, stress test, and gated SPECT characteristics of the validation group according to all-cause mortality and MACE.

|  | **Overall**<br>**(n=766)** | **Alive**<br>**(n=698)** | **All-cause**<br>**mortality**<br>**(n=68)** | ***P*-value** | **Without**<br>**MACE**<br>**(n= 694)** | **With**<br>**MACE**<br>**(n=72)** | ***P*-value** |
| --- | --- | --- | --- | --- | --- | --- | --- |
| **Demographics** |  |  |  |  |  |  |  |
| Age (years), mean (SD) | 66.9 ± 11.2 | 66.1 ± 11.2 | 74.1 ± 7.9 | < 0.001 | 66.3 ± 11.3 | 71.8 ± 7.9 | <0.001 |
| Age > 69 years, *n* (%) | 380 (49.6%) | 324 (46.4) | 56 (82.4) | <0.001 | 330 (47.6) | 50 (69.4) | 0.001 |
| Body surface (m<sup>2</sup>), mean (SD) | 1.8 ± 0.2 | 1.8 ± 0.17 | 1.8 ± 0.2 | 0.479 | 1.8 ± 0.17 | 1.8 ± 0.15 | 0.332 |
| Body mass index (kg/m<sup>2</sup>), mean (SD) | 27.5 ± 4.2 | 27.6 ± 4.3 | 26.6 ± 3.5 | 0.070 | 27.5 ± 4.3 | 26.8 ± 3.5 | 0.191 |
| Obesity (BMI ≥ 30), *n* (%) | 154 (20.1) | 145 (20.7) | 9 (13.2) | 0.139 | 142 (20.5) | 12 (16.7) | 0.444 |
| **Medical history** |  |  |  |  |  |  |  |
| Hypertension, *n* (%) | 465 (60.7) | 416 (59.6) | 49 (72.1) | 0.045 | 416 (59.9) | 49 (68.1) | 0.180 |
| Hypercholesterolemia, *n* (%) | 401 (52.4) | 368 (52.7) | 33 (48.5) | 0.509 | 367 (52.9) | 34 (47.2) | 0.360 |
| Smoking, *n* (%) | 122 (15.9) | 114 (16.3) | 8 (11.8) | 0.326 | 112 (16.1) | 10 (13.9) | 0.620 |
| Diabetes mellitus, *n* (%) | 175 (22.9) | 140 (20.1) | 35 (51.5) | < 0.001 | 139 (20) | 36 (50) | < 0.001 |
| CRF ≥ 1 (%) | 642 (83.8) | 581 (83.2) | 61 (89.7) | 0.167 | 576 (83) | 66 (91.67) | 0.057 |
| Previous MI, *n* (%) | 169 (22) | 142 (20.3) | 27 (39.7) | < 0.001 | 133 (19.2) | 36 (50) | < 0.001 |
| Previous CR, *n* (%) | 99 (12.9) | 91 (13) | 8 (11.8) | 0.765 | 90 (12.9) | 9 (12.5) | 0.910 |
| **Electrocardiogram** |  |  |  |  |  |  |  |
| Sinus rhythm, *n* (%) | 766 (100) | 698 (100) | 68 (100) | - | 694 (100) | 72 (100) | - |
| Left bundle branch block, *n* (%) | 48 (6.3) | 37 (5.3) | 11 (16.2) | < 0.001 | 38 (5.5) | 10 (13.9) | 0.005 |
| **Symptoms at gated SPECT** |  |  |  |  |  |  |  |
| Angina, *n* (%) | 101 (13.2) | 88 (12.6) | 13 (19.1) | 0.130 | 82 (11.8) | 19 (26.4) | 0.001 |
| **Medical treatment at gated SPECT** |  |  |  |  |  |  |  |
| Beta-blocker, *n* (%) | 282 (36.8) | 250 (35.8) | 32 (47.1) | 0.067 | 245 (35.3) | 37 (51.4) | 0.007 |
| Nitrate, *n* (%) | 232 (30.3) | 202 (28.9) | 30 (44.1) | 0.009 | 193 (27.8) | 39 (54.2) | < 0.001 |
| Calcium-channel blocker, *n* (%) | 126 (16.5) | 114 (16.3) | 12 (17.7) | 0.780 | 112 (16.1) | 14 (19.4) | 0.471 |
| **Stress test** |  |  |  |  |  |  |  |
| Exercise test, *n* (%) | 487 (63.6) | 452 (64.8) | 35 (4.3) | 0.030 | 445 (64.1) | 42 (58.3) | 0.331 |
| Exercise test plus pharmacological test, *n* (%) | 132 (17.2) | 123 (17.6) | 9 (13.2) | 0.361 | 123 (17.7) | 9 (12.5) | 0.264 |
| Pharmacological test, *n* (%) | 147 (19.2) | 123 (17.6) | 24 (35.3) | < 0.001 | 126 (18.2) | 21 (29.2) | 0.024 |
| Stress angina, *n* (%) | 95 (12.4) | 93 (13.3) | 2 (2.9) | 0.013 | 91 (13.1) | 4 (5.6) | 0.064 |
| ↓ ST mm ≥ 1, *n* (%) | 99 (12.9) | 90 (12.9) | 9 (13.2) | 0.936 | 84 (12.1) | 15 (20.8) | 0.036 |
| **SPECT** |  |  |  |  |  |  |  |
| SRS, mean (SD) | 1.4 ± 3.1 | 1.2 ± 3 | 3.1 ± 3 | < 0.001 | 1.2 ± 2.9 | 3.4 ± 3 | < 0.001 |
| SRS% > 9.6, *n* (%) | 45 (5.9) | 36 (5.2) | 9 (13.2) | 0.007 | 34 (4.9) | 11 (15.3) | < 0.001 |
| SSS, mean (SD) | 2.2 ± 3 | 1.9 ± 3.1 | 5.2 ± 4 | < 0.001 | 1.8 ± 3 | 5.3 ± 4 | < 0.001 |
| SSS% > 6.6, *n* (%) | 136 (17.8) | 105 (15) | 31 (45.6) | < 0.001 | 101 (14.6) | 35 (48.6) | < 0.001 |
| SDS, mean (SD) | 0.8 ± 1 | 0.7 ± 1.2 | 2.1 ± 2 | < 0.001 | 0.7 ± 1.5 | 2 ± 2 | < 0.001 |
| SDS% > 5.2, *n* (%) | 52 (6.8) | 38 (5.4) | 14 (20.6) | < 0.001 | 40 (5.8) | 12 (16.7) | < 0.001 |
| **Gated** |  |  |  |  |  |  |  |
| EDV (ml), mean (SD) | 80.1 ± 42.9 | 77.9 ± 41 | 102.3 ± 51 | < 0.001 | 78.7 ± 42.6 | 92.9 ± 44 | 0.008 |
| EDV index (ml/m<sup>2</sup>), mean (SD) | 44.6 ± 24.3 | 43.3 ± 23.5 | 57.2 ± 28.6 | < 0.001 | 43.8 ± 24 | 52.1 ± 25.2 | 0.006 |
| EDV index > 38 ml (ml/m<sup>2</sup>), *n* (%) | 358 (467) | 311 (44.6) | 47 (69.1) | < 0.001 | 313 (45.1) | 45 (62.5) | 0.005 |
| ESV (ml), mean (SD) | 36.9 ± 20 | 35 ± 30 | 57 ± 40 | < 0.001 | 35.6 ± 30 | 49.1 ± 36 | 0.003 |
| ESV index (ml/m<sup>2</sup>), mean (SD) | 20.5 ± 20 | 19.5 ± 16 | 32 ± 24 | < 0.001 | 19.7 ± 10 | 27.6 ± 20 | 0.002 |
| ESV index > 15 ml (ml/m<sup>2</sup>), *n* (%) | 342 (44.6) | 294 (42.1) | 48 (70.6) | < 0.001 | 297 (42.8) | 45 (62.5) | 0.001 |
| LVEF (%), mean (SD) | 60.5 ± 16.3 | 61.6 ± 15 | 49 ± 15.8 | < 0.001 | 61.4 ± 16 | 51.2 ± 15.9 | < 0.001 |
| LVEF < 50%, *n* (%) | 190 (24.8) | 155 (22.2) | 35 (51.5) | < 0.001 | 155 (22.3) | 35 (48.6) | < 0.001 |

Abbreviations as in Table 1.

## DISCUSSION

The importance of creating a model for coronary risk stratification in women with simple variables which are already known, lies in the following; firstly, women have often been underrepresented in cardiovascular studies^22^; secondly, cardiovascular disease is the leading cause of mortality and also the leading cause of premature death in women in some countries^27, 28^; thirdly, women are more likely than men to die in the year following an acute myocardial infarction^27, 28^; and fourthly, most women have at least one major cardiovascular risk factor (diabetes mellitus, hypertension, dyslipidemia, smoking, obesity, physical inactivity, depression, and anxiety)^27, 28^. In our total cohort [n=2226] 83.8% have ≥ 1 major cardiovascular risk factor.

The present study demonstrates the feasibility of how the risk of MACE in women can be calculated by means of CORSWO. This novel clinical risk prediction model for women used different known variables derived from clinical, stress test, and gSPECT-MPI data. This new risk score allows the risk to be calculated for individual women in a simple way with a mean of a 4-year follow-up; additionally, it includes 13 variables: age > 69 years, diabetes, left bundle branch block, angina history, treatment with nitrate, pharmacological stress, ↓ ST mm ≥ 1, stress angina, myocardial necrosis > 9.6%, myocardial ischemia > 5.2%, myocardial perfusion stress > 6.6%, ESV-index > 15 ml, and LVEF < 50%.

In our series, the most important clinical variables are: age > 69 years, diabetes, left bundle branch block, angina history, and nitrate treatment. In regards to age, in all studies, it is a prognostic variable for MACE, and women are generally older when diagnosed of coronary artery disease and have a higher prevalence of comorbid conditions^28^. It is important to note that 50% of women aged older than 75 years are diagnosed with acute myocardial infarction without chest pain^29^. In addition, diabetes was one of the most important predictors of MACE. Recently, in a prospective study of 41,350 diabetic participants without cardiovascular disease a total of 6,945 suffered cardiovascular events during the follow-up^30^. Diabetic women have a 3- to 7-fold increased risk of developing CAD^31^, and have a significantly higher mortality after myocardial infarction than diabetic men^32^. According to LBBB, most studies looking at LBBB include male patients, and the presence of LBBB is strongly associated with future high-degree atrioventricular block^33^. In the observational retrospective CODE study, 1,529,971 patients were investigated with a mean follow-up of 3.7 years, and LBBB had higher risk of overall mortality (HR 1.77; CI 95% 1.55 to 2.01)^34^. Also, in some patients, septal strain patterns evaluated by mean of echocardiography reflect the severity of LBBB induced left ventricular dysfunction^35^. Moreover, women are more likely to develop angina pectoris as their first coronary artery disease symptom and are less likely to present an acute myocardial infarction^36^. On the other hand, women often present angina-equivalent symptoms such as shortness of breath, palpitations, and fatigue^36^, and other studies have shown that the proportion of patients with myocardial infarction without chest pain is significantly higher in women than men^36^. Despite all these considerations, angina in our series has a prognostic value for MACE. Furthermore, the use of nitrate has a prognostic value, because the reason for the indication of nitrites was the presence of angina. Recently, in a large series of 5,707 patients, the use of nitrates was found to have prognosis value for non-fatal MI, CR and CD in patients with suspected CAD (HR 1.3; 95% CI: 1.05 to 1.7) or established (HR 1.4; 95% CI: 1.2 to 1.8) CAD regardless of gender^37^. Previously, in other studies, it has been observed that other clinical variables are related to cardiovascular mortality in women, for example: obesity and physical inactivity^1^, metabolic syndrome^38^, hypertension^27^, tobacco use^27^, cholesterol^27^, depression^39^, anxiety-related psychiatric disorder^40^ and menopause^41^. In our study, 86% of the women were > 55 years, probably most of the women were in the menopausal stage. Menopause is defined as cessation of menses after 12 months^11^. This typically occurs at age 51, and there is an increased risk for CAD in women 10 years after the onset of menopause^11^. This is related to our cut-off age of 69 years as a predictor of MACE. Thus, in women there are multiple clinical variables related to cardiac complications during follow-up that need to be taken into account in risk stratification.

The most important exercise and gSPECT-MPI variables included in CORSWO are: ↓ ST mm ≥ 1, stress angina, pharmacological stress, myocardial necrosis > 9.6%, myocardial perfusion stress > 6.6%, myocardial ischemia > 5.2%, ESV-index > 15 ml and FEVI < 50%. It is well known that ST depression during stress test is a criterion for myocardial ischemia and has prognostic value in women^42^. Furthermore, in the Finnish Cardiovascular Study (FINCAVAS), it was observed that the ST-segment depression/heart rate (ST/HR) hysteresis is a good method for CAD detection in women^43^. Nonetheless, in women there may be false positives, so it is more convenient to associate imaging techniques with stress tests^44^. gSPECT-MPI provides a more sensitive and specific prediction of the presence of CAD than exercise ECG does, and the reported sensitivity of exercise MPI has generally been placed in the range of 85%-90% and specificity in the range of 70%-75%^7^. Therefore, and as previously commented by Acampa et al., in women at an intermediate pretest probability of disease, a noninvasive imaging-based test for ischemia, combining exercise ECG with MPI is preferred, whenever local expertise and availability could permit^7^. Regarding to different types of stress tests (exercise, exercise plus vasodilator drugs and only vasodilator drugs), patients studied with vasodilator drugs have a worse prognosis (2.9, 95% CI: 1.7 to 4.9) for cardiac death or non-fatal MI^45^. In a long series of 6,069 patients with a mean follow-up of 10 years, those undergoing adenosine test had more mortality than the patients who underwent exercise test^46^. Besides this, the MACE with adenosine was comparable to that of patients with low exercise capacity with exercise duration of less than 3 minutes. Within the propensity-matched cohorts, the annualized MACE rate in the adenosine vs exercise patients also remained two folds higher (1.1% vs 0.2%, P<0.001)^46^. This result is comparable with other series that include 2,922 patients with a mean follow-up of 5 years since the patients underwent dipyridamole had a worse prognosis^45^. Symptomatic or asymptomatic myocardial ischemia detected in different imaging techniques^14, 15, 17, 37^ is one of the variables with the highest prognostic value known for decades. The SSS is another predictive variable with prognostic value. Previously, 2,194 women were studied with an intermediate to high CAD pre-test risk by means of gSPECT-MPI with a mean follow-up of 2.4 years. It was observed that gSPECT-MPI provides risk stratification for cardiac events (non-fatal MIs, cardiac deaths, unstable angina requiring hospitalization, and late revascularizations) beyond standard exercise stress testing for women with suspected CAD, especially in patients with intermediate Duke Treadmill Score and it is independent of estrogen status^47^. The odds ratio for SSS >3, SSS 4-8 and SSS > 8 were 2.5 (95% CI: 1.5 to 4.4), 1.9 (95% CI: 1.01 to 3.8), 4.5 (95% CI: 2 to 10) respectively. The SRS is generally an underestimated perfusion parameter, despite this it has prognostic value since it is an indicator of myocardial scarring. In the Myoview Prognosis Registry, 7,849 outpatients were enrolled^48^. In this study, it was observed that as the percent myocardium with resting defects worsen, overall CAD event rates increased regardless of gender. And on the other hand, for patients with 10% or more of the rest myocardium with perfusion defects, cardiovascular death or MI rates ranged from 7% to 44% (*P* < 0.0001)^48^. The LVEF and ESV are two recognized variables that can influence in the prognostic of patients because they are a direct expression of the systolic left ventricular function and remodeling. Therefore, in a contemporary cohort of women, we can obtain an excellent coronary risk stratification in women, however at the expense of multiple variables.

This study has limitations, first, because patients included in this study were from a single tertiary teaching hospital, indiscriminate extrapolation of the findings should be avoided; though this study presents a high number of enrolled patients with a long follow-up. Second, this study was a retrospective analysis of data, though extracted from a prospectively collected database in a reference University Hospital where the patients were meticulously studied. Lastly, of the 9,160 women, 3,525 were excluded because only had images of stress without rest. Despite this, this may not affect our results, since it is an extensive cohort with more than 2000 patients.

## Data Availability

All data is available

## NONSTANDARD ABBREVIATIONS AND NONSTANDARD ACRONYMS

HR: high risk
LR: low risk
MR: moderate risk
VHR: very high risk
VLR: very low risk

## ACKNOWLEDGMENTS

The investigators thank the women who participated in this study. The authors are grateful to Rosemay Chehab (London) for her grammatical English correction.

## SUPPLEMENTAL MATERIAL

Table S1. Figure S1, Figure S2.

